# Vitamin D-related polymorphisms and vitamin D levels as risk biomarkers of COVID-19 infection severity

**DOI:** 10.1101/2021.03.22.21254032

**Authors:** Ana Teresa Freitas, Conceição Calhau, Gonçalo Antunes, Beatriz Araújo, Matilde Bandeira, Sofia Barreira, Filipa Bazenga, Sandra Braz, Daniel Caldeira, Susana Constantino Rosa Santos, Ana Faria, Daniel Faria, Marta Fraga, Beatriz Nogueira-Garcia, Lúcia Gonçalves, Pavlo Kovalchuk, Luísa Lacerda, Hugo Lopes, Daniel Luís, Fábio Medeiros, Ana M. P. Melo, José Melo-Cristino, Ana Miranda, Clara Pereira, Ana Teresa Pinto, João Pinto, Helena Proença, Angélica Ramos, João P. R. Rato, Filipe Rocha, Júlio César Rocha, André Moreira-Rosário, Helena Vazão, Yuliya Volovetska, João-Tiago Guimarães, Fausto Pinto

## Abstract

**Background:** Vitamin D is a fundamental regulator of host defences by activating genes related to innate and adaptive immunity. Previous research shows a correlation between the levels of vitamin D in patients infected with SARS-CoV-2 and the degree of disease severity. This work investigates the impact of the genetic background related to vitamin D pathways on COVID-19 severity. For the first time, the Portuguese population was characterized regarding the prevalence of high impact variants in genes associated with the vitamin D pathways.

**Methods:** This study enrolled 517 patients admitted to two tertiary Portuguese hospitals. The serum concentration of 25 (OH)D, was measured in the hospital at the time of patient admission. Genetic variants, 18 variants, in the genes *AMDHD1, CYP2R1, CYP24A1, DHCR7, GC, SEC23A*, and *VDR* were analysed.

**Results:** The results show that polymorphisms in the vitamin D binding protein encoded by the *GC* gene are related to the infection severity (*p* = 0.005). There is an association between vitamin D polygenic risk score and the serum concentration of 25 (OH)D (*p* = 0.042). There is an association between 25 (OH)D levels and the survival and fatal outcomes (*p* = 1.5e-4). The Portuguese population has a higher prevalence of the *DHCR7* RS12785878 variant when compared with its prevalence in the European population (19% versus 10%).

**Conclusion:** This study shows a genetic susceptibility for vitamin D deficiency that might explain higher severity degrees in COVID-19 patients. These results reinforce the relevance of personalized strategies in the context of viral diseases.

**Trial registration:** NCT04370808

## Introduction

From the end of 2019 until now, a new coronavirus named SARS-CoV-2 has been causing a worldwide pandemic leading to many deaths due to an acute respiratory infection named COVID-19. The lack of knowledge about the pathogenesis of this disease and the reasons underlying the different clinical outcomes have been pushing the society as an all to take preventive measures and to accelerate research. During the last year, several research groups and healthcare institutions have made available evidence that indicates that an uncontrolled inflammatory response is a major responsible for the occurrence of an acute respiratory distress syndrome (1).

A statistical analysis of COVID-19 pandemic data from healthcare centers across China, France, Germany, Italy, Iran, South Korea, Spain, Switzerland, the United Kingdom and the United States, performed by Northwestern University, shows a strong correlation between severe vitamin D deficiency and mortality rates (2).

Vitamin D has many relevant and documented roles in general health maintenance, being its deficiency particularly associated with severe impacts on the functional integrity of the immune system, such as influencing cytokine production (3). Vitamin D deficiency has also been linked to hypertension, autoimmune, infectious and cardiovascular diseases, also known risk factors for severe COVID-19 (4).

Previous research shows that genetics contribute to up to 28% of inter-individual variability in serum 25(OH)D concentrations, while season and vitamin D intake explain another 24% of the variability (5). Large-scale genome-wide association studies (GWAS), considering 79,366 individuals with European ancestry (6), or considering imputed genotypes from 401,460 white British UK Biobank participants (7), have identified relatively common single nucleotide polymorphisms which play an important biological role in vitamin D metabolism, transport, degradation and downstream pathways, to evaluate their impact on circulating 25(OH) D concentrations. Vitamin D polymorphisms research shows that not all genomes will respond to vitamin D supplementation due to the loss of function of different genes (8).

In a normal scenario, it is known that vitamin D deficiency is common in Europe and the Middle East. It occurs in <20% of the population in Northern Europe, in 30–60% in Western, Southern and Eastern Europe and up to 80% in Middle East countries (9). Data from Portugal shows that 66% of adults present vitamin D insufficiency/deficiency (10). This high percentage in a population living in a sunny country, motivated the characterization that was performed regarding the prevalence of high impact genetic variants in genes associated with the vitamin D pathways in the Portuguese population when compared with the European population.

The main objective of this study was to understand if an association exists between polymorphisms in vitamin D-related genes and vitamin D levels, and between these variables and the COVID-19 severity. Secondarily, we aimed to compare the frequency of the genetic variants under analysis with the observed frequency for the European population. To achieve this objective, we have assessed the genetic variants in vitamin D-related genes in hospitalized patients and the vitamin D serum levels, and evaluated the association between these data and the severity of the disease.

## Results and discussion

A total of 491 patients with a laboratory confirmed positive COVID-19 test, 371 (75.6%) from Santa Maria hospital and 120 (24.4%) from São João hospital, were considered. There were 217 female and 266 male patients with mean ± SD age of 69.7±15.8 years (see supplemental data for demographic, clinical and phenotypic data).

In COVID-19 positive patients, 311 (63.3%), 120 (24.4%) and 59 (12.0%) patients had insufficient, deficient and sufficient levels of vitamin D, respectively, using the Endocrine Society cutoff (see supplemental data). The prevalence of vitamin D deficiency was 61.7% and 68.3% in Santa Maria and São João hospitals, respectively.

Dead, severe and moderate disease were observed in 18.5%, 21.8% and 59.7% of patients, respectively. From a total of 311 patients with vitamin D deficiency, 68 died (13.8% of all patients), 69 (14% of all patients) had a severe response and 174 (35.4 % of all patients) had a moderate response to COVID-19. Disease severity for each patient was defined using the World Health Organization (WHO) discrete clinical progression scale from 0 to 10 (11). In this study patients’ severity levels were between 4 and 10.

### Identification of the vitamin D polymorphisms as risk biomarkers

From previous large-scale GWAS, several single nucleotide polymorphisms that play an important biological role in vitamin D metabolism, transport, degradation, and downstream pathways, have been identified as having an impact on circulating 25(OH) D concentrations (6, 7). To understand if an association exists between the polymorphisms in the vitamin D-related genes and the disease severity, four polygenic risk scores (PRSs) were defined. These scores considered contributions from different genes and were identified as: Synthesis (*DHCR7; CYP2R1*); Metabolism (*GC; CYP24A1*); Pathway (*DHCR7; CYP2R1; GC; CYP24A1*) and Vitamin D total (*DHCR7; CYP2R1; GC; CYP24A1; AMDHD1; SEC23A*).

Table 1 represents the correlations between the PRSs and COVID-19 disease severity, and the PRSs and vitamin D levels. The results show a significant positive correlation between the Metabolism score and COVID-19 disease severity, and a significant negative correlation between the Vitamin D total score and vitamin D levels.

**Table 1.**
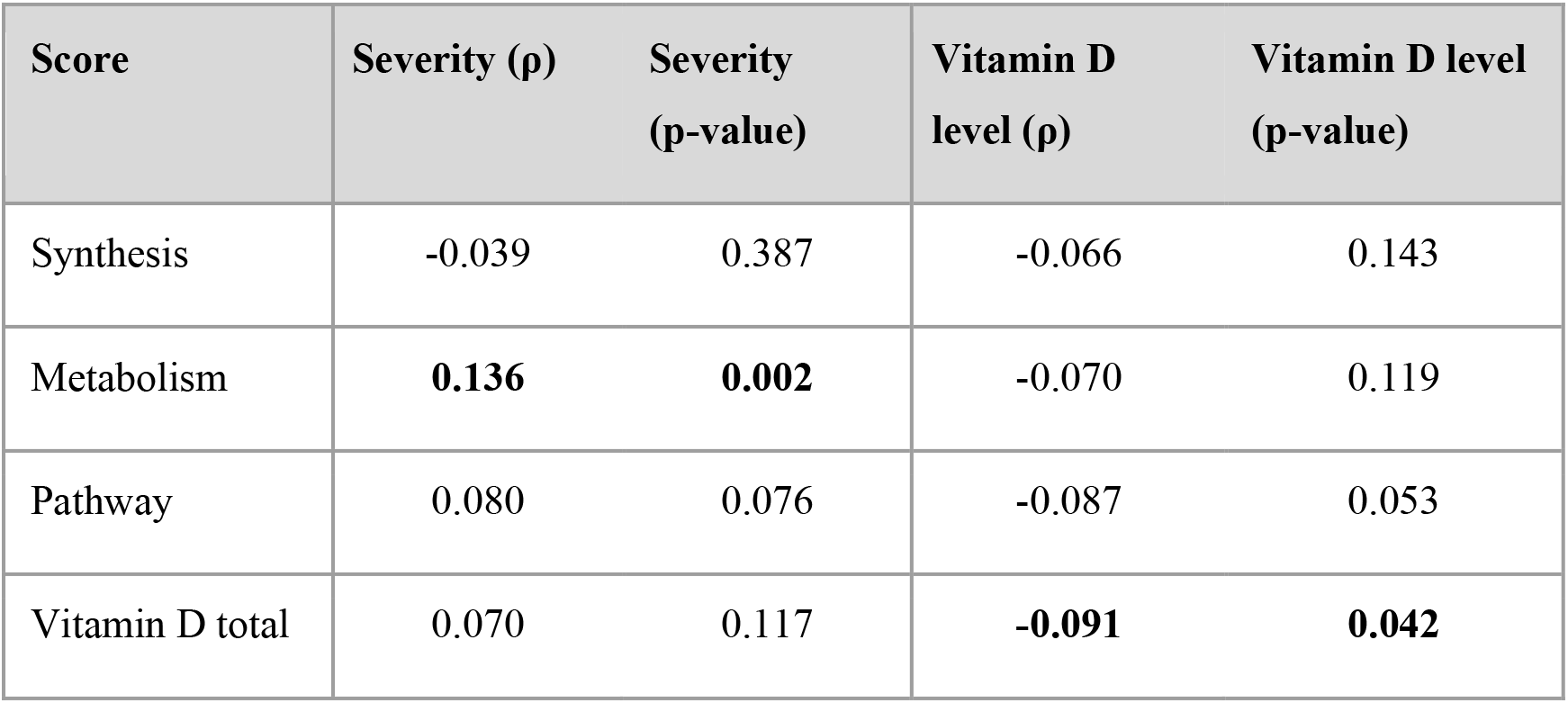
PRSs correlation with COVID-19 disease severity and vitamin D level (Spearman correlation).

The relation between the Vitamin D total score values and vitamin D levels can be clearly verified in Figure 1. Higher PRSs values are associated with patients with deficient levels of vitamin D, with a higher aggregation of points at the median/high level of the PRSs in vitamin D deficient patients.

**Figure 1.**
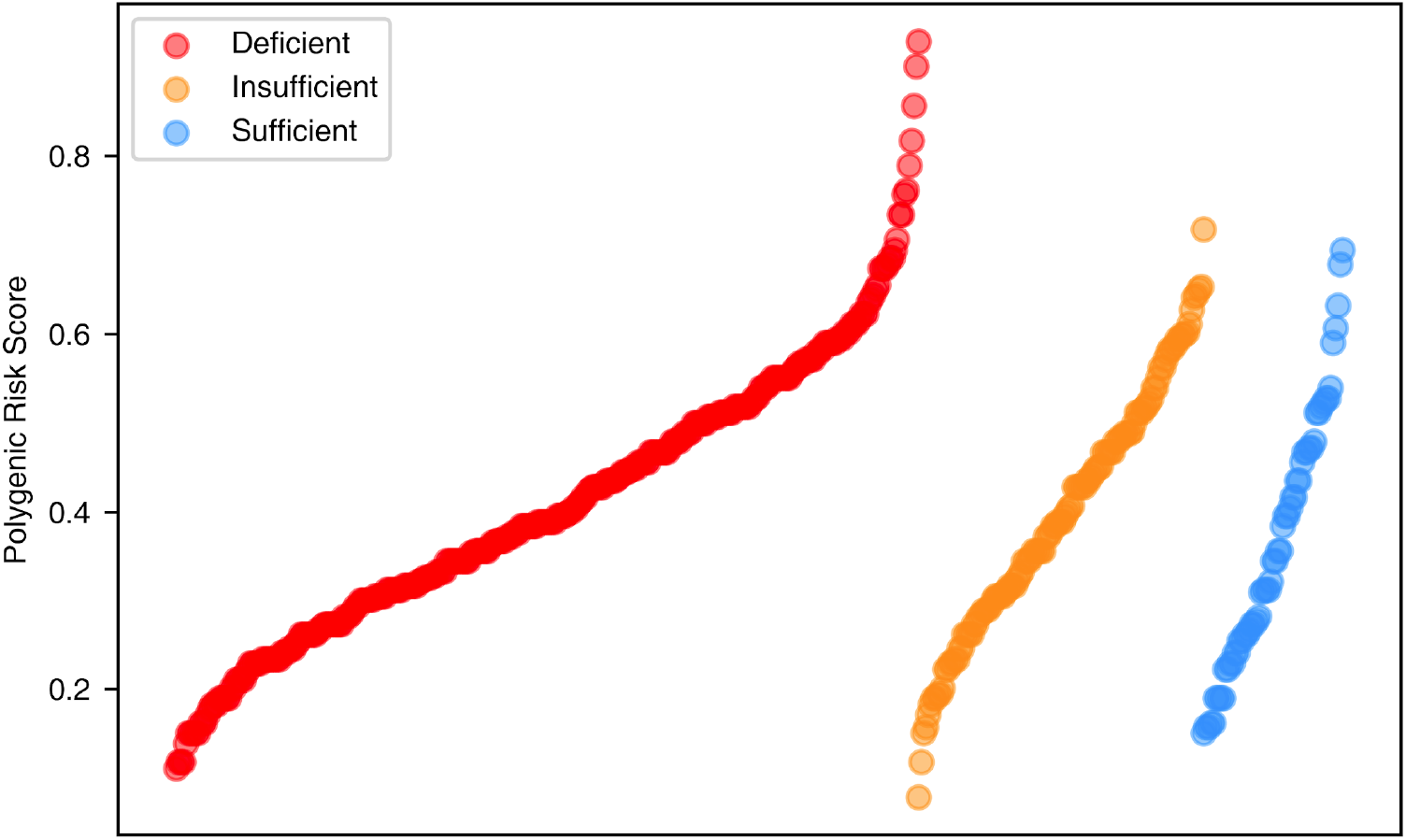
PRSs are represented as a continuous value. Patients were divided in three groups (Deficient - red; Insufficient - yellow; Sufficient - blue) considering its vitamin D level (a continuous variable). In each group, patients are sorted in ascending order of PRSs.

A deeper analysis of the positive correlation between the Metabolism score and COVID-19 disease severity showed that the polymorphism *GC* RS2282679 in the vitamin D binding protein encoded by the *GC* gene could explain most of the interesting correlation observed. The association of this polymorphism with the severity presents a ρ = 0.13 (p-value = 0.005).

This finding was particularly relevant because only one previous GWAS meta-analysis from the COVID-19 host genetics initiative considering hospitalized vs. not hospitalized COVID-19 patients (https://www.covid19hg.org/results/) briefly highlighted this fact. In this meta-analysis, the *GC* RS2282679 correlated significantly with COVID-19 disease severity (p-value = 0.002).

Regarding the association of the *GC* RS2282679 polymorphism with the disease severity, it might be explained by an additional role played by the vitamin D binding protein other than the transport of vitamin D in the bloodstream. Available data indicates that this protein may also act as a neutrophil chemotactic factor and a macrophage activator, therefore actively participating in the inflammation process (12, 13, 14). Also, vitamin D binding protein is an extracellular scavenger for actin released from damaged/dead cells. When in excess, actin can cause intravascular coagulation resulting in multi-organ dysfunction and cardiac arrest (15).

The functional consequence of this polymorphism in this pathway needs to be further explored in future studies.

### Relation between vitamin D level and COVID-19 disease severity

It was observed a trend towards an increased proportion of patients with deficient levels of vitamin D (< 20 ng/ml) and an increase of severity from moderate to severe, and dead. The proportion of patients with deficient levels of vitamin D was higher in the group that died (76%) when compared with the two other groups (moderate - 59% and severe - 64% disease). The combined proportion of patients with insufficient and sufficient vitamin D levels was 40% in the group with moderate severity, compared with 24% in the group of deceased patients (see supplemental material).

This evidence was also confirmed when a binomial variable of survival and fatal outcome was used (Figure 2). Vitamin D levels were significantly lower in patients that died than in those that survived (Median = 11.70 [Q1 = 8.67, Q3 = 19.67] vs Median = 17.40 [Q1 = 11.00, Q3 = 24.60], respectively (p-value = 1.5e-4, Mann-Whitney U test). These results corroborate what was already observed by other studies across the world.

**Figure 2.**
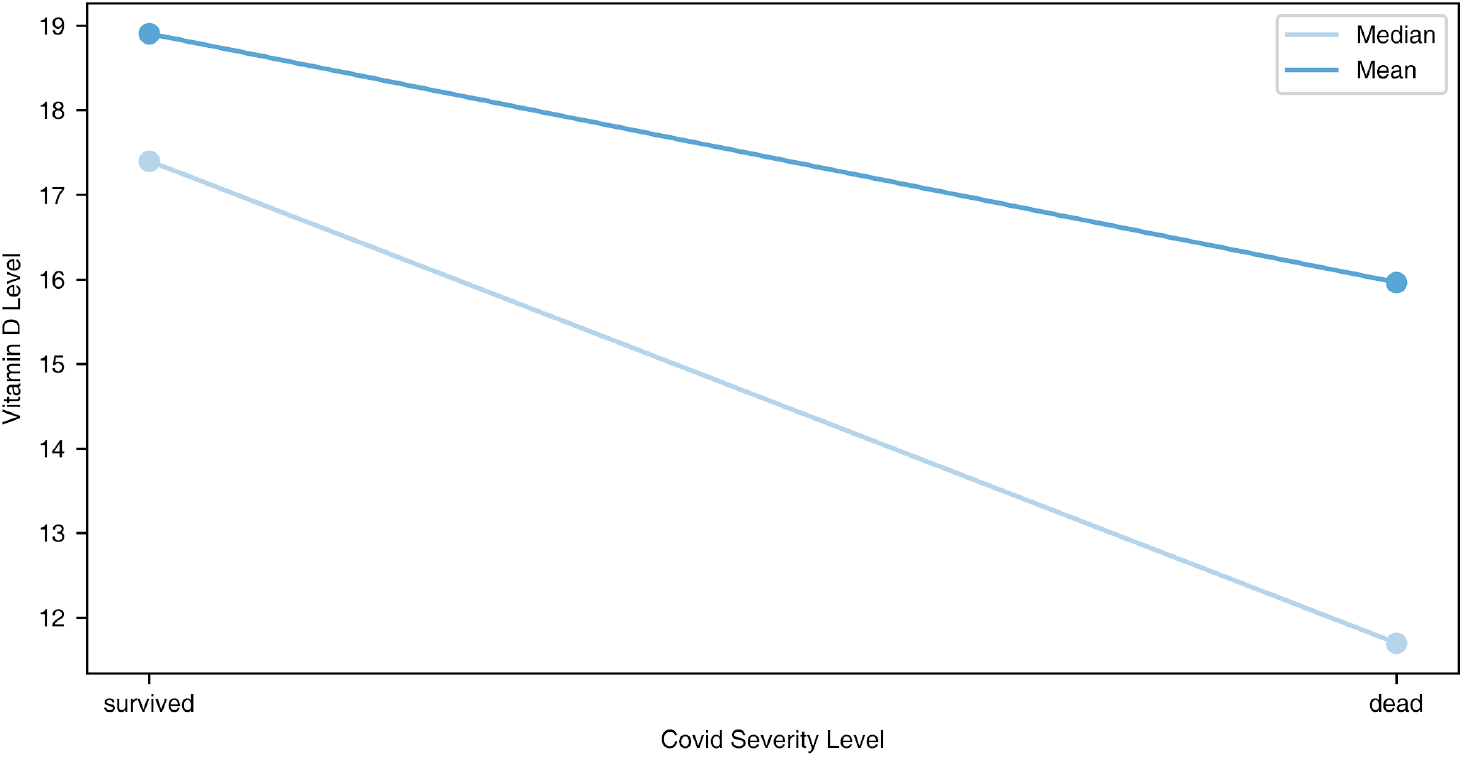
Vitamin D level (ng/mL) (continuous) and COVID-19 disease severity (binomial: survived, dead)

### Differences between impact genotypes’ frequency in Portuguese and European populations

The genotypes’ frequency of the genetic variants under analysis was compared with the observed frequency for the European population in the 1000 Genomes Project, Figure 3. The variant *DHCR7* RS12785878 shows a significant deviation in the impact genotypes’ frequency (G/GT), 18.9% for the Portuguese population compared to 9.7% for the European population. The prevalence of the impact polymorphism *DHCR7* RS12785878 is similar in the Portuguese population in the HeartGenetics’s research database with more than 8,000 Portuguese individuals (data not shown), where GG genotype has a prevalence of 18.8%.

**Figure 3.**
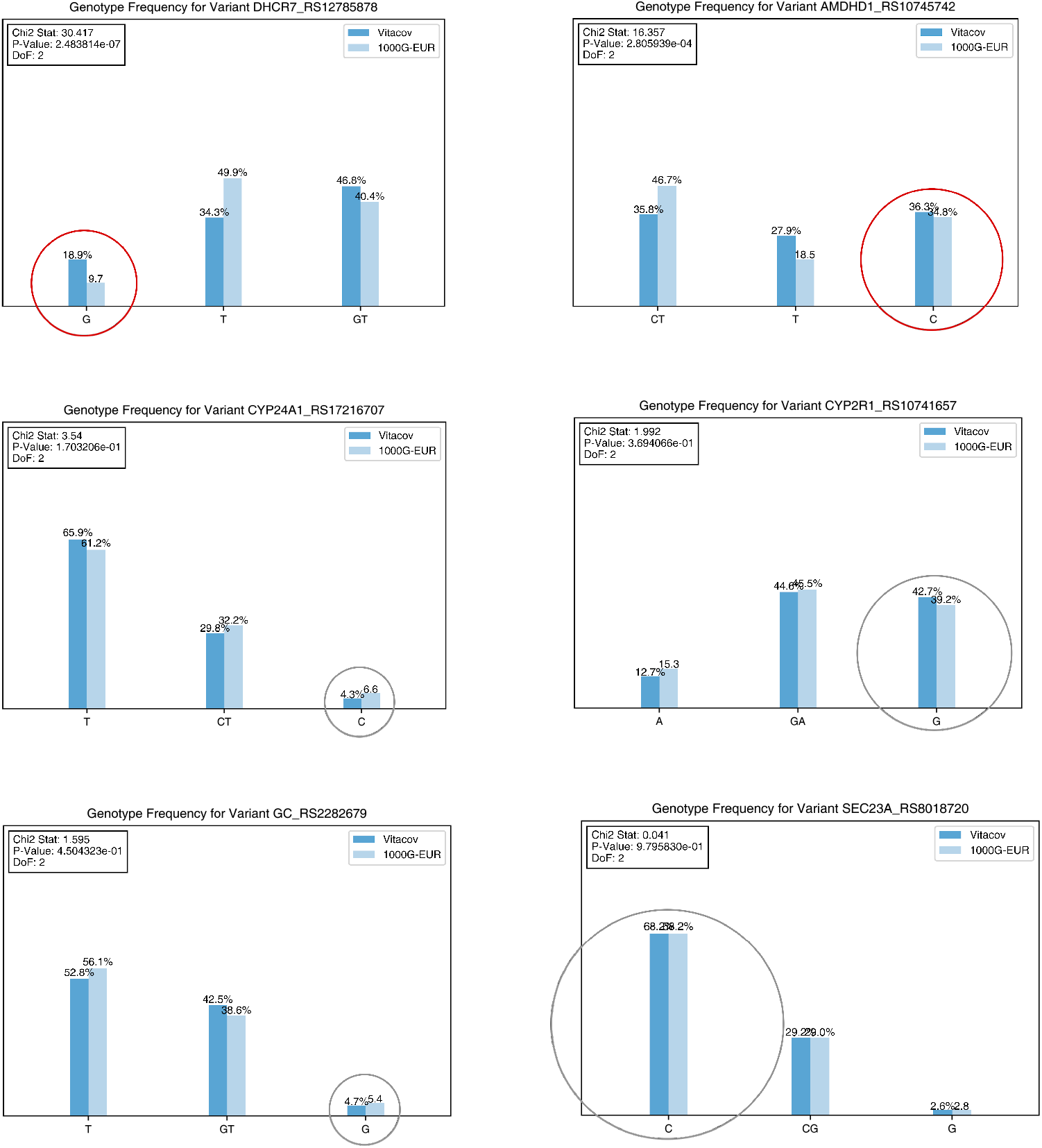
Comparison of the impact genotypes’ frequency in the Portuguese and European populations. Circles highlight the risk genotype. The red circles show the risk genotypes from variants with a statistically significant difference between these two populations (p-value = 2.5e-7 for *DHCR7* RS12785878, and p-value = 2.8e-4 for *AMDHD1* RS10745742).

In this study, and for the first time, it is shown that the Portuguese population presents a genetic makeup for a higher predisposition to vitamin D deficiency when compared to the European population, therefore increasing the potential severity of the COVID-19 response, consequently impacting on patient outcomes. Furthermore, these results also explain, in part, the higher vitamin D deficiency in Portuguese population in different previous publications (10). It is important to know that this metabolic vulnerability must be considered for a vitamin D supplementation clinical decision. This data proves that it is wrong to assume that sunny countries would not have an issue with vitamin D deficiency, showing that genetic characterization and vitamin D monitoring at a population level should be put in place in order to define guidelines for vitamin D intake.

One year after this pandemic scenario, it has been described that vitamin D deficiency may be a risk factor for mortality in COVID-19 infected patients (9). On top of this, evidence revealed that supplementation with high-dose vitamin D3 booster therapy and reduced the risk of mortality (16). Whereas vitamin D intake, sun exposure, demographics, and, specially, genes have been identified as being crucial determinants of vitamin D status, the impact of these factors is expected to be different across populations. To improve current prevention and treatment strategies, it is essential to propose novel diagnostic tools for personalization.

These results reinforce the role of vitamin D polymorphisms, in particular the *GC* RS2282679 polymorphism, and vitamin D levels as biomarkers for COVID-19 disease severity and emphasize the relevance for personalized strategies in the context of viral diseases. Whether these relationships are causal or consequential is unknown. Nevertheless, they represent targets for diagnostic surveillance, or for intervention studies.

## Methods

### Subjects

Between August 2020 and January 2021, 517 patients were enrolled in this project when admitted to Hospital de Santa Maria in Lisbon and Hospital de São João in Oporto.

Eligibility criteria included patients aged 18 years and above, patients admitted with COVID-19 disease requiring hospitalization, with a positive test for SARS-CoV-2 by nasopharyngeal swabs using quantitative RT-PCR performed in national reference laboratories and in accordance with recommendations from the National Directorate of Health. All eligible patients were followed till the closure of case i.e. either discharge or mortality.

### Serum 25-Hydroxyvitamin

*D*. Vitamin D status was determined by analyzing 25(OH) D concentrations in the serum samples collected at the time of patient admission. Serum 25(OH) D was determined in the local pathology laboratories by a competitive electrochemiluminescence protein-binding immunoassay using a Cobas® e411 or e801 automated analyzer (Roche Diagnostics GmbH). This assay uses a vitamin D-binding protein as capture protein, which binds to both forms of vitamin D: 25(OH) D2 and 25(OH) D3. Thus, in this report, the terms ‘vitamin D’ and ‘25(OH) D’ refer to both forms: 25(OH) D2 and 25(OH) D3. The specific terms for vitamin D2 and D3 are used to refer to the corresponding individual form. The 25(OH) D cut-offs established by the Clinical Guidelines Subcommittee of the Endocrine Society were followed, considering the absence of a global consensus on the 25(OH) D concentration that defines vitamin D deficiency and “adequate” 25(OH) D levels for extra-skeletal functions (17). Thus, vitamin D adequacy was classified according to the following 25(OH) D cut-off levels: deficiency, <20 ng/mL; insufficiency, 20–29 ng/mL; and sufficiency, ≥30 ng/mL (18). The season of blood sample collection was also considered, as it may affect the patient’s vitamin D concentrations (19).

### Genotyping

All patients signed an informed consent to perform a genetic test for the analysis of 18 genetic variants that play a role in vitamin D metabolism, transport, degradation and downstream pathways. See supplemental data for details about the genetic panel, Table ii. The genetic test was performed using the iPLEX® MassARRAY® system at the HeartGenetics’ certified laboratory in Portugal. DNA samples were prepared from blood samples by HeartGenetics, CCUL and São João hospital laboratories.

### Data management

Clinical history, genotypic and phenotypic data, stored at the e-CRF, were collected and managed using REDCap electronic data capture tools (20) hosted by BioData.pt (https://biodata.pt/), the Portuguese distributed infrastructure for biological data, at INESC-ID research institute. All datasets are pseudo-anonymous and only one of them has a key that connects to the patient. In accordance with the GDPR, only the PI of the project (Prof. Fausto Pinto) and the clinicians responsible for data acquisition have access to this key. All project participants have controlled data access. The clinical history and the patient informed consent were supervised by the Cardiology Service at Santa Maria hospital and by the Pathology Service at São João hospital (see supplemental data for e-CRF content details).

### Data analysis

Clinical history, genotypic and phenotypic data was evaluated using statistical, machine learning and polygenic risk scores methodologies. Vitamin D polymorphisms prevalence in different populations was obtained from 1000 Genomes database (https://www.internationalgenome.org/) and from HeartGenetics’s research database for the Portuguese population, with more than 8,000 Portuguese individuals.

Regarding the methodological approach, the following steps were undertaken:

1. *Data cleaning and validation:* All variables were analysed for outliers and missing values. Some discrepancies, such as different units of measure and data entry errors, were identified and fixed. No imputation was made. Regarding data transformation, both disease severity and vitamin D levels were categorized in different levels, and the genetic variants were aggregated in PRSs.
2. *Descriptive analysis:* A complete, graphical descriptive analysis of the data was created for all variables of interest as univariate analysis. Data are presented as numbers or percentages for categorical variables, while continuous variables are shown as mean and standard deviation, and median and interquartile range (25th percentile - 75th percentile).
3. *Analysis of data distribution:* The data normality was accessed using Shapiro-Wilk test and D’Agostino Pearson’s test. Statistical normality testing is relevant in order to set up the category of statistical methods (parametric or non-parametric) used in further analysis. Results showed that most parameters do not follow a normal distribution, thus for further analysis it was considered only non-parametric statistical tests that do not assume any particular data distribution.
4. *Identification of the vitamin D polymorphisms as risk biomarkers:* Several statistical tests were used, namely Mann-Whitney and Kruskal-Wallis Tests. Spearman rank correlation coefficient was also calculated. Four PRSs have been computed, focused on the vitamin D metabolism, transport and degradation pathways, based on an additive weighted model, having values in the interval [0, 1]. In this interval, 0 corresponds to a lower risk of having low vitamin D levels due to genetics, and 1 corresponds to a higher risk of having low vitamin D levels due to genetics (see supplemental material for details about the PRSs). The four different scores considered the following genetic variants.
  1. Synthesis score = *DHCR7* RS12785878 + *CYP2R1* RS10741657
  2. Metabolism score = *GC* RS 2282679 + *CYP24A1* RS17216707
  3. Pathway score = *DHCR7* RS12785878 + *CYP2R1* RS10741657 + *GC* RS 2282679 + *CYP24A1* RS17216707
  4. Vitamin D total score = *DHCR7* RS12785878 + *CYP2R1* RS10741657 + *GC* RS 2282679 + *CYP24A1* RS17216707 + *AMDHD1* RS10745742 + *SEC23A* RS8018720
  The PRSs did not model other genetic variants that have been tested, since their impact has not been obtained by the same GWAS studies, which could introduce a bias in its relative impact.
5. *Analysis of the correlation between hypovitaminosis D and the disease severity:* Different statistical tests were employed, namely Mann-Whitney and Kruskal-Wallis Tests, depending on the type of categorization under analysis. Spearman rank correlation coefficient was also calculated in order to analysed not only an eventual association but also to quantify it and observe its direction.
6. *Genotypes frequency comparison:* For this comparison the 1000 Genomes (https://www.ensembl.org/index.html) and the HeartGenetics’s research database with more than 8,000 Portuguese individuals were used.

Concerning the different statistical tests performed, a p-value < 0.05 was considered statistically significant.

### Study approval

This project was approved by the Ethics Committee of Hospital de Santa Maria in Lisbon and Hospital de São João in Oporto. The study was conducted in accordance with the ethical principles of the Declaration of Helsinki and followed the Good Clinical Practice guidelines. Written informed consent was obtained from all study participants prior to their inclusion in the study.

## Data Availability

For the moment the data used in this project is just available to all participant institutions.
Clinical history, genotypic and phenotypic data, stored at the e-CRF, were collected and managed using REDCap electronic data capture tools hosted by BioData.pt (https://biodata.pt/), the Portuguese distributed infrastructure for biological data, at INESC-ID research institute. All datasets are pseudo-anonymous and only one of them has a key that connects to the patient. In accordance with the GDPR, only the PI of the project (Prof. Fausto Pinto) and the clinicians responsible for data acquisition have access to this key. All project participants have controlled data access. The clinical history and the patient informed consent were supervised by the Cardiology Service at Santa Maria hospital and by the Pathology Service at Sao Joao hospital.

## Author contributions

ATF conceived and supervised the study, analysed the results, and wrote the manuscript with input from all the authors. MB, SB, SB, DC, SCRS, MF, BNG, FM, JMC, AM, ATP, HP, FR, YV and FP were responsible for sample and data collection and interpretation from 370 patients of Hospital Santa Maria. BA, FB, LL, JP, AR and JTG were responsible for sample and data collection and interpretation from 120 patients of Hospital São João. HV, LG and CP were responsible for genetic testing and sample logistics and performed the patients genotyping. GA supervised the study and the bioinformatics team. HL and PK performed the bioinformatics analyses. DL designed the genetic testing panel and analysed the results. DF, JPRR and AMPM developed the e-CRM and performed data cleaning. AF, JCR contributed to the interpretation of the analysis. AM-R contributed to statistical analysis, and data interpretation. CC, JTG and FP conceived and supervised the study, and analysed the results. All authors revised and approved the manuscript.

## Acknowledgments

We thank all the study participants, who donated blood and authorized the genetic analysis and the collection of the clinical and phenotypic personal data. This project was supported by the “Fundação para a Ciência e Tecnologia”, program “Research 4 Covid-19 Apoio especial a projetos de implementação rápida para soluções inovadoras de resposta à pandemia de COVID-19”.

