## Supplemental data for "Vitamin D-related polymorphisms and vitamin D levels as risk biomarkers of COVID-19 infection severity"

1 Centro Cardiovascular da Universidade de Lisboa (CCUL), CAML, Faculdade de Medicina, Universidade de Lisboa, Lisboa, Portugal;
2 HeartGenetics, Genetics and Biotechnology SA, Biocant Park, Portugal;
3 INESC-ID, Instituto Superior Técnico, University of Lisbon, Lisboa, Portugal;
4 Nutrition and Metabolism, NOVA Medical School, Universidade Nova de Lisboa, Lisboa, Portugal;
5 CINTESIS - Center for Health Technology and Services Research, Portugal;
6 Rheumatology Department, Hospital de Santa Maria, Centro Hospitalar Universitário Lisboa Norte, Centro Académico de Medicina de Lisboa, Lisboa, Portugal;
7 Internal Medicine Department, Santa Maria Hospital, Centro Hospitalar Universitário Lisboa Norte, Faculdade de Medicina da Universidade de Lisboa, Portugal.
8 Department of Infectious disease, Santa Maria Hospital - CHULN, Lisboa, Portugal
9 Clinical Pathology Department, Hospital Santa Maria, Centro Hospitalar Universitário Lisboa Norte, EPE, Lisbon, Portugal;
10 Cardiology Department, Hospital Universitário de Santa Maria - CHULN, Lisboa, Portugal;
11 Serviço de Patologia Clínica, Centro Hospitalar Universitário de São João
12 Departamento de Biomedicina, Faculdade de Medicina, EPIUnit, Instituto de Saúde Pública, Universidade do Porto;
13 Portuguese Infrastructure of Biological Data – BioData.pt;
14 Instituto Gulbenkian de Ciência, Oeiras, Portugal.
15 Unidade de Investigação em Reumatologia, Instituto de Medicina Molecular, Faculdade de Medicina, Universidade de Lisboa, Centro Académico de Medicina de Lisboa, Lisboa, Portugal

### Supplemental data

#### Demographic, clinical and phenotypic characteristics

For the data analysis, a total of 491 patients with a laboratory confirmed positive COVID-19 test and completed data registries, 371 (75.6%) from Santa Maria hospital and 120 (24.4%) from São João hospital, were considered. There were 217 female and 266 male patients with COVID-19 with mean ± SD age of 69.7±15.8 years. Dead, severe and moderate disease were observed in 18.5%,

21.8% and 59.7% of patients, respectively. Demographic and clinical characteristics of these patients are reported in Table i below.

The majority of individuals had three or more pre-existing comorbidities, with hypertension (63.1%), diabetes (31.8%) and obesity (23.4%) diseases being the most frequent ones (Figure i).

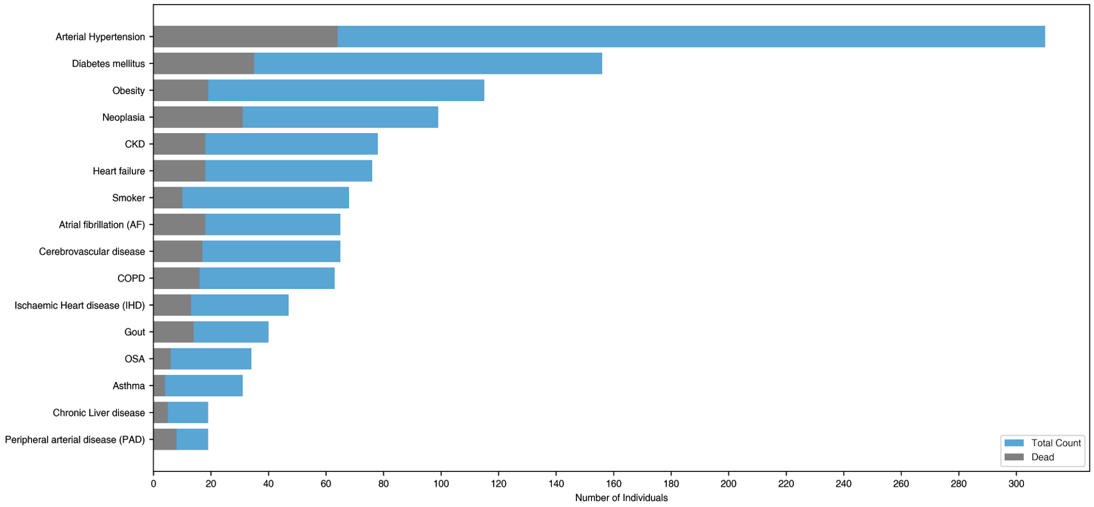


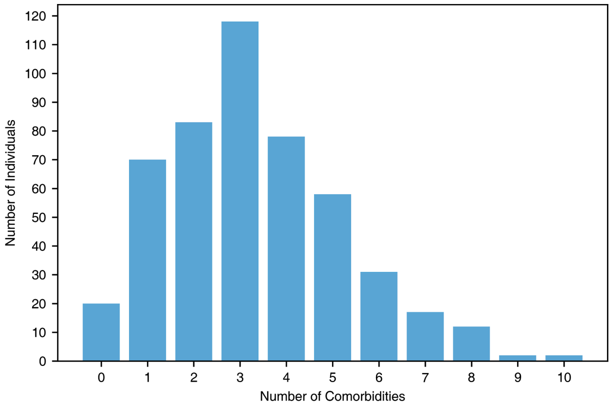


Figure i: Frequency of comorbidities (a), and distribution of the number of significant comorbidities (b).

During hospitalization, most patients needed oxygen supply (76.4%), and 22.2 percent were admitted to the ICU due to the necessity of non-invasive or invasive mechanical ventilation as determined by the health care providers. The mean length of stay was 17.9±16.7 days, and 91 patients died.

In COVID-19 positive patients, the prevalence of vitamin D deficiency was 61.7% and 68.3% in Santa Maria and São João hospitals, respectively (Figure ii), using the Endocrine Society cutoff (Figure iii). On Chi square test, the differences in the prevalence of vitamin D deficiency (vitamin D level < 20 ng/mL), insufficient (vitamin D level [20, 30[ ng/mL) and sufficient (vitamin D level >= 30 ng/mL), among the two hospitals, were 6.6%, 11.4% and 3.9%, respectively. These differences are statistically significant with a p-value of 0.036.  The analysis related with the Vitamin D levels was also performed for each hospital's dataset separately. It was observed that the correlation results pointed in the same direction in both subsets and gained statistical significance when they were combined in a single dataset (when compared with each independent result). The reduced sample size of each hospital's subset leads to less statistical power in the results, particularly in São João's dataset that is smaller. Considering these observations, the combined dataset was chosen to be the reference for the presented results.

From a total of 311 patients with vitamin D deficiency, 68 died, 69 had a severe response and 174 had a moderate response to COVID-19.

Table i: Demographic and clinical characteristics of the patients

| | | # | % |
|---|---|---|---|
| Patients | | 491 | 100.0 |
| Age | | 69.7±15.8 | - |
| Sex | Male | 266 | 54.2 |
| | Female | 217 | 44.2 |
| | n.a. | 8 | 1.6 |
| COVID-19 severity (WHO clinical progression scale - Table ii) | 4 | 110 | 22.4 |
| | 5 | 183 | 37.3 |
| | 6 | 77 | 15.7 |
| | 7 | 6 | 1.2 |
| | 8 | 12 | 2.4 |
| | 9 | 12 | 2.4 |
| | 10 | 91 | 18.5 |

| | | | |
|---|---|---|---|
| Vitamin D levels | deficient | 311 | 63.3 |
| | insufficient | 120 | 24.4 |
| | sufficient | 59 | 12.0 |
| Comorbidities | Arterial hypertension | 310 | 63.1 |
| | Diabetes mellitus | 156 | 31.8 |
| | Obesity | 115 | 23.4 |
| | Neoplasia | 99 | 20.2 |
| | CKD | 78 | 15.9 |
| | Heart failure | 76 | 15.5 |
| | Smoker | 68 | 13.8 |
| | Atrial fibrillation (AF) | 65 | 13.2 |
| | Cerebrovascular disease | 65 | 13.2 |
| | COPD | 63 | 12.8 |
| | Ischaemic heart disease (IHD) | 47 | 9.6 |
| | Gout | 40 | 8.1 |
| | OSA | 34 | 6.9 |
| | Asthma | 31 | 6.3 |
| | Chronic liver disease | 19 | 3.9 |
| | Peripheral arterial disease | 19 | 3.9 |
| Drugs | Statins | 184 | 37.5 |
| | Diuretic | 166 | 33.8 |
| | Beta-blocker | 133 | 27.1 |
| | Calcium channel blockers | 125 | 25.5 |
| | ARA | 121 | 24.6 |
| | Anti-Aggregate | 104 | 21.2 |
| | ACE inhibitors | 96 | 19.6 |
| | Metformin | 93 | 18.9 |
| | Anticoagulant | 68 | 13.8 |

| | | | |
|---|---|---|---|
| | DPP-4 inhibitors | 49 | 10.0 |
| | Corticosteroids | 46 | 9.4 |
| | Insulin | 38 | 7.7 |
| | Vitamin D medication | 33 | 6.7 |
| | Non-steroidal anti-inflammatory | 28 | 5.7 |
| | ADO - others | 26 | 5.3 |
| | Immunosuppressant - others | 24 | 4.9 |
| | Spironolactone | 20 | 4.1 |
| | iSGLT2/aGLP1 | 20 | 4.1 |
| | Antiarrhythmics | 16 | 3.3 |

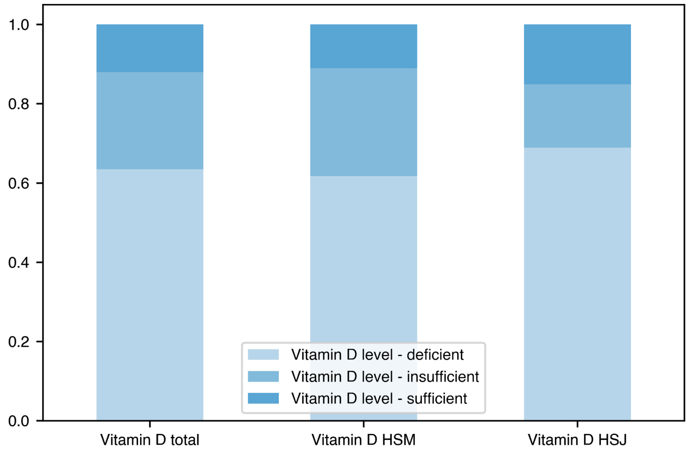


Figure ii: Vitamin D levels (categories %) in all recruited patients and by hospital distribution. (Deficient: < 20 ng/mL; Insufficient: [20, 30[ ng/mL; Sufficient: >= 30 ng/mL).

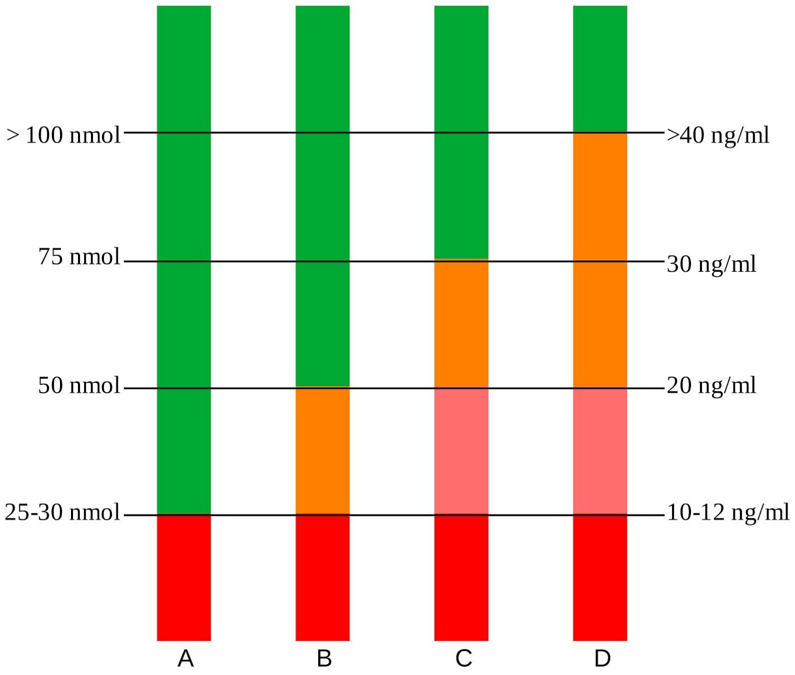


Figure iii: Interpretation of serum values of 25 (OH)D according to different agencies and countries. red- severe deficiency; orange – mild deficiency; green – sufficient supply. A: Scientific Advisory Committee on Nutrition; Netherlands. B: Institute of Medicine; Australia-New Zealand; Nordic and Deutschland (Germany), Austria and Confoederatio Helvetica (Switzerland) countries; American Academy of Pediatrics. C: Endocrine Society; International Osteoporosis Foundation; American Geriatrics Society. D: Vitamin D Council and a 'few experts'. Adapted from (1).

In the data analysis the vitamin D levels were evaluated as a continuous variable (ng/ml) and as a categorical variable, following the Endocrine Society guideline: Deficient: < 20 ng/ml; Insufficient: [20, 30[ ng/ml; Sufficient: >= 30 ng/ml.

The following table presents the World Health Organization (WHO) clinical progression scale that was used to define the patient disease severity. The data analysis was performed considering the following values for the variable COVID-19 disease severity:

- {4, 5, 6, 7, 8, 9, 10} (ordinal) - there are no uninfected or mild disease cases in this study.
- {Moderate disease, Severe disease, Dead} (categorical)
- {Survived, Dead} (binomial)

Table iii: WHO clinical progression scale for patient disease severity. Adapted from (4).

| **Patient State** | **Descriptor** | **Score** |
|---|---|---|
| Uninfected | Uninfected, no viral RNA detected | 0 |
| Ambulatory mild disease | Asymptomatic; viral RNA detected | 1 |
| | Symptomatic; independent | 2 |
| | Symptomatic; assistance needed | 3 |
| Hospitalised; moderate disease | Hospitalised; no oxygen therapy | 4 |
| | Hospitalised; oxygen by mask or nasal prongs | 5 |
| Hospitalised: severe disease | Hospitalised; no oxygen by NIV or high flow | 6 |
| | Intubation and mechanical ventilation, $pO_2/FiO_2 \geq 150$ or $SpO_2/FiO_2 \geq 200$ | 7 |
| | Mechanical ventilation $pO_2/FiO_2 < 150$ ($SpO_2/FiO_2 < 200$) or vasopressors | 8 |
| | Mechanical ventilation $pO_2/FiO_2 < 150$ and vasopressors, dialysis, or ECMO | 9 |
| Dead | Dead | 10 |

### Genetic panel

Table ii describes the genes list and genetic variants that have been tested for each patient. Each gene is described with the following information: gene name, polymorphism RS code, information about the encoded protein, the impact allele for the decreased 25(OH)D, the variant type.

Table ii: Genetic parameters.

| **Vitamin D Pathway** | | **Encoded protein** | **Effect allele for decreased 25(OH)D** | **Variant type** |
|---|---|---|---|---|
| CYP2R1 | rs10741657 | Vitamin D 25-hydroxylase | G (major) | 5' UTR |
| CYP2R1 | rs12794714 | | A (minor) | Synonymous (NP_078790.2:p.Ser59=) |
| CYP2R1 | rs7116978 | | C (major) | Intron Variant |
| GC | rs2282679 | Vitamin D-binding protein | G (minor) | Intron Variant |

| | | | | |
|---|---|---|---|---|
| GC | rs1155563 | | C (minor) | Intron Variant |
| GC | rs7041 | | A (minor) | Missense (NP_001191236.1: p.Asp451Glu) |
| DHCR7 | rs12785878 | 7-Dehydrocholesterol Reductase | G (minor) | Downstream (NADSYN1 : Intron Variant) |
| DHCR7 | rs12800438 | | G (minor) | Downstream (NADSYN1 : Intron Variant) |
| DHCR7 | rs4944957 | | A (minor) | Downstream (NADSYN1 : Intron Variant) |
| CYP24A1 | rs6013897 | Vitamin D(3) 24-Hydroxylase | A (minor) | Upstream (intergenic) |
| CYP24A1 | rs17216707 | | C (minor) | Upstream (intergenic) |
| CYP24A1 | rs6127099 | | T (minor) | Upstream (intergenic) |
| AMDHD1 | rs10745742 | Amidohydrolase Domain Containing 1 | C (major) | Intron Variant |
| SEC23A | rs8018720 | Protein Transport Protein Sec23A [promotes the formation of transport vesicles from the endoplasmic reticulum (ER)] | C (major) | Missense (NP_006355.2:p.Leu211Val (G)) Missense (NP_006355.2:p.Leu211Ile (A)) |
| VDR | rs7975232 | Vitamin D receptor | NA | Intron Variant |
| VDR | rs1544410 | | NA | Intron Variant |
| VDR | rs2228570 | | NA | Missense (NP_001017535.1:p.Met1Thr) |
| VDR | rs731236 | | NA | Synonymous (NP_001017535.1:p.Ile352=) |

### Clinical and phenotypic parameters stored at the e-CRF

Table iii describes the e-CRF main statistics. Each clinical and phenotypic parameters and corresponding information is presented in Table iv.

Clinical history, genotypic and phenotypic data, stored at the e-CRF, was collected and managed using REDCap electronic data capture tools hosted by BioData.pt (https://biodata.pt/), the Portuguese distributed infrastructure for biological data, at INESC-ID research institute. REDCap (Research Electronic Data Capture) is a secure, web-based software platform designed to support data capture for research studies, providing 1) an intuitive interface for validated data capture; 2) audit trails for tracking data manipulation and export procedures; 3) automated export procedures for seamless data downloads to common statistical packages; and 4) procedures for data integration and interoperability with external sources (5, 6). All datasets are pseudo-anonymous and only one of them has a key that connects to the patient.

Table iii: e-CRF data description: main statistics.

| | |
|---|---|
| # patients | 517 |
| # clinical parameters | 91 |
| # genetic parameters | 18 |
| # complete records | 491 |

Table iv: Clinical and phenotypic parameters.

| Section | Variable | Variable Type |
|---|---|---|
| **Patient identification** | Review of inclusion and exclusion criteria | Nominal qualitative variables |
| **Clinical and Demographics** | Age | Discrete quantitative |
| | Weight | Continuous quantitative |
| | Height | Continuous quantitative |
| | BMI | Continuous quantitative |
| | Sex | Nominal qualitative |
| | Symptoms | Nominal qualitative |
| | Co-morbidities | Nominal qualitative |
| **Hospitalization** | Medication | Nominal qualitative variables |
| | Admission Analysis | Nominal quantitative variables |

| | | |
|---|---|---|
| | Transferred to the Intensive Care Unit during hospitalization? | Nominal qualitative variables |
| | Oxygen by mask or nasal prongs | |
| | Oxygen by NIV or high-flow | |
| | Intubation and mechanical ventilation with pO2/FiO2 >= 150 or SpO2/FiO2 >= 200 | |
| | Mechanical ventilation with pO2/FiO2 < 150 or SpO2/FiO2 < 200, or use of vasopressors | |
| **Outcomes** | Mechanical ventilation with pO2/FiO2 < 150 or SpO2/FiO2 < 200, and use of vasopressors, dialysis or ECMO | |
| | Dead | |
| | EAM | |
| | TEP | |
| | Stroke | |
| | Discharged up to 60 days after admission? | |
| | Length of hospitalization | Continuous quantitative |

**Polygenic Risk Score**

A Polygenic Risk Score (PRS) is an estimate of an individual's genetic liability to a trait or disease, calculated according to their genotype profile and relevant GWAS data.

For a set of genetic variants x, with weights β,

(1) $x = \{x_1, \ldots, x_n\}$

(2) $\beta = \{\beta_1, \ldots, \beta_n\}$

Given an impact function g(x) that returns an impact value for a certain genetic variant x,

(3)

$$g(x) = \begin{cases} 1 & \text{if } x \text{ is impact genotype} \\ 0.5 & \text{if } x \text{ is heterozygous} \\ 0 & \text{otherwise} \end{cases}$$

The PRS is calculated by the following function $f_\beta(x)$,

$$(4) \qquad f_\beta(x) = \frac{\sum_{i=1}^{n} \beta_i \cdot g(x_i)}{\sum_{i=1}^{n} \beta_i}$$

For this project, the considered genetic variants and the corresponding impact alleles were selected from GWAS studies conducted in cohorts ranging from 33 996 to 443 374 European individuals showing reproducible genomic hits associated with variation in serum 25(OH)D (2, 3).

The following table describes the values used for the parameter β, for six polymorphisms. These values, which represent the impact of each polymorphism in the model, were obtained simultaneously by a GWAS study with 79,366 individuals with European ancestry (2).

Table iv: Parameters used in the PRS.

| **Gene** | **rsID** | **Effect allele for decreased 25(OH)D** | $\beta$ | **p-value** |
|---|---|---|---|---|
| CYP2R1 | rs10741657 | G (major) | -0.031 | 2.05e-46 |
| GC | rs2282679 | G (minor) | -0.089 | 4.74e-343 |
| DHCR7 | rs12785878 | G (minor) | -0.036 | 3.80e-62 |
| CYP24A1 | rs17216707 | C (minor) | -0.026 | 8.14e-23 |
| AMDHD1 | rs10745742 | C (major) | -0.017 | 1.88e-14 |
| SEC23A | rs8018720 | C (major) | -0.017 | 4.72e-9 |

The PRSs did not model other genetic variants that have been tested, since their impact has not been obtained by the same GWAS studies, which could introduce a bias in its relative impact. Simulations have been performed considering the impact of *VDR* gene polymorphisms but no association improvement with COVID-19 severity was observed.

**Methodological approach**

Regarding the methodological approach, the following steps were undertaken:

1. *Data cleaning and validation:* all variables were analyzed for outliers and missing values. Some discrepancies, such as different units of measure and data entry errors, were identified and fixed. No imputation was made. Regarding data transformation, both disease severity and vitamin D levels were categorized in different levels, and the genetic variants were aggregated in PRSs.

2. *Descriptive analysis:* a complete, graphical descriptive analysis of the data was created for all variables of interest as univariate analysis. Data are presented as numbers or percentages for categorical variables, while continuous variables are shown as mean and standard deviation, and median and interquartile range (25th percentile - 75th percentile).

3. *Analysis of data distribution:* This step provides a clear understanding of what is the underlying distribution that the analyzed parameters follow in the dataset. Statistical normality testing is relevant in order to set up the category of statistical methods (parametric or non-parametric) used in further analysis. The data normality was accessed using Shapiro-Wilk test and D'Agostino Pearson's test. Results showed that most parameters do not follow a normal distribution, thus for further analysis it was considered only non-parametric statistical tests that do not assume any particular data distribution.

4. *Identification of the vitamin D polymorphisms as risk biomarkers:* This step focused on finding differences in genetic variants in vitamin D-related genes between COVID-19 patients with different degrees of disease severity. Several statistical tests were used (following the same assumptions in the former topic), namely Mann-Whitney and Kruskal-Wallis Tests. Spearman rank correlation coefficient was also calculated.
   Four PRSs have been computed, focused on the vitamin D metabolism, transport and degradation pathways, based on an additive weighted model, having values in the interval [0, 1]. In this interval, 0 corresponds to a lower risk of having low vitamin D levels due to genetics, and 1 corresponds to a higher risk of having low vitamin D levels due to genetics (see supplemental material for details about the PRSs). The four different scores considered the contribution of the following genetic variants.

   (1) Synthesis score = *DHCR7* RS12785878 + *CYP2R1* RS10741657

(2) Metabolism score = *GC* RS 2282679 + *CYP24A1* RS17216707

(3) Pathway score = *DHCR7* RS12785878 + *CYP2R1* RS10741657 + *GC* RS 2282679 + *CYP24A1* RS17216707

(4) Vitamin D total score = *DHCR7* RS12785878 + *CYP2R1* RS10741657 + *GC* RS 2282679 + *CYP24A1* RS17216707 + *AMDHD1* RS10745742 + *SEC23A* RS8018720

5. *Analysis of the correlation between hypovitaminosis D and the disease severity:* This step focused on finding differences in vitamin D blood levels between COVID-19 patients with different degrees of disease severity. Different statistical tests were employed, namely Mann-Whitney and Kruskal-Wallis Tests, depending on the type of categorization under analysis. Spearman rank correlation coefficient was also calculated in order to analyze not only an eventual association but also to quantify it and observe its direction.

6. *Genotypes frequency comparison:* For this comparison the 1000 Genomes (https://www.ensembl.org/index.html) and the HeartGenetics's research database with more than 8,000 Portuguese individuals were used.

Concerning the different statistical tests performed, a p-value $< 0.05$ was considered statistically significant.
